# Comparative effectiveness of bivalent BA.4.5 or BA.1 mRNA booster vaccines among immunocompromised individuals across three Nordic countries: a nationwide cohort study

**DOI:** 10.1101/2024.05.02.24306733

**Authors:** Mie Agermose Gram, Emilia Myrup Thiesson, Nicklas Pihlström, Jori Perälä, Eero Poukka, Tuija Leino, Rickard Ljung, Niklas Worm Andersson, Anders Hviid

**Author notes:** Corresponding author: Mie Agermose Gram, Department of Epidemiology Research, Statens Serum Institut, Artillerivej 5, 2300 Copenhagen S, Denmark.

## Abstract

**Objectives:** To estimate the effectiveness and waning immunity of the bivalent BA.4-5 or BA.1 mRNA booster vaccine against Covid-19-related hospital admission and death in immunocompromised individuals.

**Design:** Nationwide cohort analyses using a matched cohort design.

**Setting:** Denmark, Finland, and Sweden, from 1 September 2022 to 31 October 2023.

**Participants:** All individuals aged 18 years or above with medical history of at least one immunocompromised condition, residency in Denmark, Finland or Sweden, no history of Covid-19-related hospitalization, and receipt of at least three Covid-19 vaccine doses as of study start, 1 September 2022. Individuals boosted with a BA.4-5 or BA.1 vaccine were matched 1:1 with unboosted individuals.

**Main outcome measures:** Country-combined vaccine effectiveness (VE) estimates against Covid-19 hospitalization and Covid-19- related death at day 270 of follow-up. Potential waning was assessed in 45-day intervals.

**Results:** A total of 352,762 BA.4-5 and 191,070 BA.1 booster vaccine doses were administered to immunocompromised individuals. At day 270, the comparative VE against Covid-19 hospitalization was 34.2% (95% CI, 7.1% to 61.3%) for the bivalent BA.4-5 vaccine (696 vs 1,128 events, risk difference [RD] per 100,000, -223.7, 95% CI, -411.5 to -36.0) and 42.6% (95% CI, 31.3% to 53.9%) for the BA.1 vaccine (395 vs 740 events, RD per 100,000, -385.0, -673.4 to -96.6) compared with matched unboosted. The comparative VE against Covid-19 death was 53.9% (95% CI, 38.6% to 69.3%) for the bivalent BA.4-5 vaccine (203 vs 457 events, RD per 100,000, -138.7, 95% CI, -195.5 to -81.9) and 57.9% (95% CI, 48.5% to 67.4%) for the BA.1 vaccine (112 vs 302 events, RD per 100,000, -220.6, -275.9 to -165.4). The VE estimates were highest in the first 45 days since eight days after vaccination (52.8% and 72.8% for bivalent BA.4-5 vaccine against Covid-19-related hospitalization and death, and 62.2% and 84.2% for bivalent BA.1 vaccine) and waned gradually during the 270 days of follow-up.

**Conclusions:** In immunocompromised individuals, vaccination with a bivalent BA.4-5 or BA.1 booster lowered the risk of Covid-19-related hospitalization and death over a follow-up period of 9 months. The effectiveness was highest during the first months since vaccination with subsequent gradual waning.

**Summary box:** *What is already known on this topic:*

- Bivalent BA.4-5 or BA.1 booster vaccination increases protection against severe Covid-19 outcomes in the general population.
- Lower effectiveness of the original monovalent Covid-19 vaccines among immunocompromised individuals has been observed relative to the effectiveness within the general population.

*What this study adds:*

- Bivalent BA.4-5 or BA.1 booster vaccination increased the protection against Covid-19 outcomes among immunocompromised individuals.
- At day 270 of follow-up, the bivalent BA.4-5 booster had prevented 223.7 hospitalizations and 138.7 deaths related to Covid-19 per 100,000 boosted individuals. For the bivalent BA.1 booster, corresponding numbers were 385.0 and 220.6, respectively.
- The vaccine effectiveness was highest during the first 45 days since eight days after vaccination (52.8% and 72.8% for bivalent BA.4-5 vaccine against Covid-19-related hospitalization and death, and 62.2% and 84.2% for bivalent BA.1 vaccine) and waned gradually during the 270 days of follow-up.

## Introduction

In the Nordic countries, the bivalent BA.4-5 and BA.1 mRNA booster vaccines were offered as a Covid-19 booster dose to target groups (the general population aged ≥60 years in Finland and ≥50 years in Denmark and Sweden, as well as immunocompromised individuals) during autumn 2022. Current evidence demonstrates that bivalent booster vaccination for the general population provides additional protection against severe Covid-19 outcomes such as Covid-19-related hospitalization and death (1-4). However, few studies have evaluated the vaccine effectiveness (VE) of the bivalent BA.4-5 and BA.1 boosters in immunocompromised individuals (5, 6). This includes lack of long-term follow-up enabling a better understanding of waning which is critical when optimizing the timing of additional boosters in this high-risk population. Additionally, previous studies suggest lower effectiveness of the original monovalent Covid-19 vaccines among immunocompromised individuals relative to that observed in the general population (7, 8). Across the three Nordic countries of Denmark, Finland, and Sweden, we estimated the effectiveness of the bivalent BA.4-5 or BA.1 mRNA booster vaccine against Covid-19 hospitalization and Covid-19-related death with up to 9 months follow-up in nationwide cohorts of immunocompromised adults aged ≥18 years.

## Methods

### Data sources

Danish, Finnish, and Swedish nationwide demography and healthcare registries were used to obtain individual-level information on exposure (Covid-19 vaccination status), outcomes (Covid-19 hospitalization and Covid-19-related death), and covariates (age, sex, region of residency, comorbidities, Covid-19 vaccine priority group, and time since SARS-CoV-2 infection, Table S2-S3). The individual-level register data were linked using the country-specific unique identifiers assigned to all residents. The study period was from 1 September 2022 to 31 October 2023.

### Study cohort

Within each country, we constructed a cohort of all immunocompromised adults on the basis of the following eligibility criteria: aged 18 years or above, medical history of at least one immunocompromised condition, residency in Denmark, Finland or Sweden, no history of Covid-19-related hospitalization, and received at least three Covid-19 vaccine doses as of study start, 1 September 2022 (the bivalent boosters were offered from September 2022 and onwards in each country). Individuals were defined as having an immunocompromised condition if having a history of either I) solid malignancy, II) hematologic malignancy, III) rheumatologic or inflammatory disorder, IV) other intrinsic immune condition or immunodeficiency, or V) organ or stem cell transplant or received a Covid-19 vaccine dose equivalent to a booster dose for immunocompromised (Table S1). We also used prior Covid-19 vaccination pattern to define immunocompromised. If individuals had any of the following Covid-19 vaccination schedule histories they were classified as immunocompromised: Either 1) any booster dose (≥3rd dose) within 90 days of the last dose, 2) receipt of a fourth dose before the official starting date of the roll-out of the 4th dose boosters for the general population, or 3) receipt of five or more vaccine doses prior to start of study period (Table S1).

### Study design

We used a matched study design to evaluate the effectiveness of a bivalent BA.4-5 or BA.1 mRNA-booster vaccine (administered as a ≥fourth dose) in comparison with being unboosted (vaccinated with at least three original monovalent Covid-19 vaccine doses). Immunocompromised individuals, who during the study period received a bivalent booster dose, were matched on the day of vaccination (index date) in 1:1 pairs with immunocompromised individuals who had received the same number of original monovalent Covid-19 vaccine doses prior to study start but had not received a bivalent booster vaccine up until that day (the index date). Individuals were matched on age (5-year bins), calendar month of their most recent original monovalent Covid-19 vaccine dose prior to study start (monthly bins), and a propensity score including sex, region of residence, comorbidities (chronic pulmonary disease, cardiovascular conditions or diabetes, autoimmunity-related conditions, cancer, and moderate to severe renal disease) (9, 10), Covid-19 vaccine priority group (severe Covid-19 risk group, healthcare personnel, or others), and time since SARS-CoV-2 infection (no previous infection, <6months since a positive PCR test result, 6-12 months, or >12 months). If individuals who were included as matched unboosted immunocompromised individuals received a bivalent booster dose later than the assigned index date, the follow-up of the pair was censored and the now vaccinated control was allowed to potentially re-enter as a boosted individual in a new matched pair on that given date.

### Outcomes

The outcomes of interest were Covid-19 hospitalization and Covid-19-related death. Covid-19 hospitalization was defined as the first inpatient hospitalization with a registered Covid-19-related diagnosis and a positive PCR test for SARS-CoV-2 (within 14 days before to 2 days after the day of admission). Covid-19-related death was defined as any death within 30 days after a positive PCR test for SARS-CoV-2. Information about data sources and country-specific definitions are available in the supplementary (Table S2).

### Statistical analysis

We matched, without replacement, on age and calendar month of last mutual Covid-19 vaccine and on the propensity scores with a caliper width of 0.01 (on the propensity score scale). We used logistic regression to estimate the propensity score of receiving a BA.4-5 or BA.1-booster dose given covariates (sex, region of residence, comorbidities, Covid-19 vaccine priority group, and time since SARS-CoV-2 infection) as predictors.

Individuals were followed from day eight after the date of the bivalent booster vaccination of the boosted individual in the matched pair (to ensure full immunization among boosted individuals) until the outcome under study, vaccination with an additional booster, death, emigration, 270 days after start of follow-up, or end of the study period, whichever occurred first.

Cumulative incidences of the outcome of interest were estimated by the Aalen-Johansen estimator with all cause death as a competing risk. Risk ratios and risk differences (RD) were calculated using the cumulative incidence at day 270 of follow-up; the comparative VE was calculated as 1 – risk ratio. The corresponding 95% confidence interval (CI) was calculated using the delta method. Country-specific estimates were combined by random-effects meta-analyses implemented using the *mixmeta* package in R (11); all reported effectiveness estimates are country-combined. Meta-analyzed RD and VE estimates can deviate when countries have few events and large variability, in consequence also producing wide 95% CI.

Waning of the comparative VE against the outcome of interest was estimated BA.4-5 and BA.1 using meta-regression. The comparative VE was estimated in 45-day intervals since eight days after bivalent booster vaccination (<45, 46-90, 91-135, 136-180, 181-225, and 226-270 days), which were then fitted to a linear regression where the slope coefficient represented the percentage point change in the comparative VE per 45 days since eight days after bivalent booster vaccination.

## Results

### Characteristics of the study population

Table 1 presents the baseline characteristics of the study cohort before and after matching. A total of 543,832 bivalent booster doses were given during the study period for the study cohort individuals before matching. Among the 543,832 booster doses, 352,762 (65%) were bivalent BA.4-5 booster doses and 191,070 (35%) were bivalent BA.1 booster doses given as a ≥fourth dose. Solid malignancy was the most common immunocompromised condition for both BA.4-5 (46%) and BA.1 (47%) boosted as well as for bivalent unboosted (45%) individuals, followed by rheumatologic or inflammatory disorder (25%, 26%, and 25%, respectively).

**Table 1.** Cohort characteristics before and after matching for the estimation of effectiveness of bivalent BA.4-5 or BA.1 mRNA booster vaccination as a ≥fourth vaccine dose in immunocompromised individuals in three Nordic countries. Values are numbers (percentages) unless stated otherwise.

|  | Before matching |  |  | After matching |  |  |  |
| --- | --- | --- | --- | --- | --- | --- | --- |
|  | BA.4-5 boosted<sup>a</sup> | BA.1 boosted<sup>a</sup> | Unboosted<sup>a</sup> | ≥Fourth dose BA.4-5 vs unboosted |  | ≥Fourth dose BA.1 vs unboosted |  |
|  |  |  |  | BA.4-5 boosted | Unboosted | BA.1 boosted | Unboosted |
| Total | 352,762 | 191,070 | 1,934,638 | 275,222 | 275,222 | 170,104 | 170,104 |
| Denmark | 125,524 | 76,186 | 605,990 | 83,566 | 83,566 | 65,799 | 65,799 |
| Finland | 76,868 | 32,054 | 461,544 | 65,852 | 65,852 | 29,372 | 29,372 |
| Sweden | 150,370 | 82,830 | 867,104 | 125,804 | 125,804 | 74,933 | 74,933 |
| **Bivalent booster vaccine dose<sup>b</sup>** |  |  |  |  |  |  |  |
| Fourth dose | 141,660 | 91,487 | 857,366 | 103,721 | 103,721 | 80,126 | 80,126 |
| Fifth dose | 141,620 | 95,515 | 663,313 | 112,606 | 112,606 | 86,184 | 86,184 |
| ≥Sixth dose | 69,482 | 4,068 | 413,959 | 58,895 | 58,895 | 3,794 | 3,794 |
| Mean age (SD), yrs | 72 (12.2) | 73 (11.5) | 71 (13.5) | 71 (12.7) | 71 (12.7) | 72 (11.5) | 72 (11.6) |
| Female sex | 185,812 (52.7) | 100,213 (52.4) | 1,015,648 (52.5) | 145,019 (52.7) | 144,794 (52.6) | 89,578 (52.7) | 88,785 (52.2) |
| Severe covid-19 risk group | 109,767 (31.1) | 51,946 (27.2) | 624,286 (32.3) | 85,442 (31.0) | 87,775 (31.9) | 44,502 (26.2) | 45,891 (27.0) |
| Healthcare workers | 15,264 (4.3) | 7,827 (4.1) | 82,387 (4.3) | 12,331 (4.5) | 12,522 (4.5) | 7,580 (4.5) | 7,394 (4.3) |
| **Immunocompromised condition** |  |  |  |  |  |  |  |
| Solid malignancy | 160,581 (45.5) | 90,018 (47.1) | 868,136 (44.9) | 124,082 (45.1) | 124,743 (45.3) | 79,360 (46.7) | 78,924 (46.4) |
| Hematologic malignancy | 29,124 (8.3) | 16,952 (8.9) | 156,663 (8.1) | 21,105 (7.7) | 21,447 (7.8) | 14,452 (8.5) | 15,425 (9.1) |
| Rheumatologic or inflammatory disorder | 88,415 (25.1) | 48,830 (25.6) | 484,534 (25.0) | 70,527 (25.6) | 69,380 (25.2) | 44,403 (26.1) | 42,791 (25.2) |
| Other intrinsic immune condition or immunodeficiency | 14,545 (4.1) | 7,678 (4.0) | 85,729 (4.4) | 11,872 (4.3) | 11,733 (4.3) | 7,023 (4.1) | 6,981 (4.1) |
| Organ or stem cell transplant | 6,726 (1.9) | 3,282 (1.7) | 39,030 (2.0) | 5,280 (1.9) | 5,160 (1.9) | 2,941 (1.7) | 3,283 (1.9) |
| ≥2 of abovementioned conditions | 19,145 (5.4) | 10,584 (5.5) | 106,477 (5.5) | 14,335 (5.2) | 14,425 (5.2) | 9,195 (5.4) | 10,035 (5.9) |
| Received a Covid-19 vaccine dose equivalent of booster dose for immunocompromised | 34,226 (9.7) | 13,726 (7.2) | 194,069 (10.0) | 28,021 (10.2) | 28,334 (10.3) | 12,730 (7.5) | 12,665 (7.4) |
| **Comorbidities** |  |  |  |  |  |  |  |
| Autoimmune related conditions | 98,996 (28.1) | 54,685 (28.6) | 543,739 (28.1) | 78,471 (28.5) | 77,145 (28.0) | 49,342 (29.0) | 48,154 (28.3) |
| Cancer | 196,271 (55.6) | 109,980 (57.6) | 1,064,080 (55.0) | 150,146 (54.6) | 151,554 (55.1) | 96,446 (56.7) | 97,847 (57.5) |
| Chronic pulmonary disease | 23,615 (6.7) | 13,614 (7.1) | 130,135 (6.7) | 18,126 (6.6) | 17,666 (6.4) | 11,746 (6.9) | 11,331 (6.7) |
| Cardiovascular condition or diabetes | 70,843 (20.1) | 39,595 (20.7) | 386,878 (20.0) | 53,502 (19.4) | 55,266 (20.1) | 33,769 (19.9) | 34,330 (20.2) |
| Renal disease | 18,314 (5.2) | 9,835 (5.1) | 106,769 (5.5) | 13,575 (4.9) | 14,358 (5.2) | 8,409 (4.9) | 9,352 (5.5) |
| **Previous PCR-confirmed SARS-CoV-2** |  |  |  |  |  |  |  |

|  | Before matching |  |  | After matching |  |  |  |
| --- | --- | --- | --- | --- | --- | --- | --- |
|  |  |  |  | ≥Fourth dose BA.4-5 vs unboosted |  | ≥Fourth dose BA.1 vs unboosted |  |
|  | BA.4-5 boosted<sup>a</sup> | BA.1 boosted<sup>a</sup> | Unboosted<sup>a</sup> | BA.4-5 boosted | Unboosted | BA.1 boosted | Unboosted |
| **infection** |  |  |  |  |  |  |  |
| <6 mths | 29,110 (8.3) | 15,608 (8.2) | 155,393 (8.0) | 22,204 (8.1) | 19,512 (7.1) | 13,873 (8.2) | 12,889 (7.6) |
| 6-12 mths | 31,164 (8.8) | 16,131 (8.4) | 153,756 (7.9) | 23,128 (8.4) | 20,674 (7.5) | 14,529 (8.5) | 13,030 (7.7) |
| >12 mths | 8,889 (2.5) | 4,777 (2.5) | 53,663 (2.8) | 7,351 (2.7) | 8,088 (2.9) | 4,437 (2.6) | 4,816 (2.8) |

<sup>a</sup>Unboosted before matching refers to unboosted individuals eligible for matching as of start of study period, and boosted before matching refers to individuals boosted with a bivalent BA.4-5 or BA.1 booster as a ≥fourth dose during study period eligible for matching. <sup>b</sup>For the unboosted columns, numbers represent the number of eligible individuals for matching or matched individuals, unboosted with the respective dose number. As individuals could contribute with both unboosted and boosted person-time during the study period (in separate matched pairs; see subsection 9.8 for details on procedures), overlap of individuals across the bivalent boosted columns with the unboosted column exists, and thus, the columns cannot be summed to the total study populations.

### Effectiveness of bivalent booster

After matching, the two cohorts comprised 275,222 (with a mean age of 71, SD 12.7 years) and 170,104 (mean age of 72, SD 11.5 years) matched pairs for the comparisons of bivalent BA.4-5 and BA.1 boosted vs unboosted, respectively. Overall, the characteristics among the matched pairs were similar to those of the pre-matched cohort and generally well-balanced with difference in proportions <10% (except for region in Finland; Figure S1-S2).

Figures 1-2 show the cumulative incidences of Covid-19-related hospitalization and death during the 270 days of follow-up, respectively, comparing BA.4-5 or BA.1 boosted with unboosted across all three Nordic countries. The cumulative incidences of the outcomes were lower for BA.4-5 or BA.1 boosted individuals than for unboosted individuals. Differences were less pronounced for Covid-19-related hospitalization in Finland after approximately 110 days of follow-up.

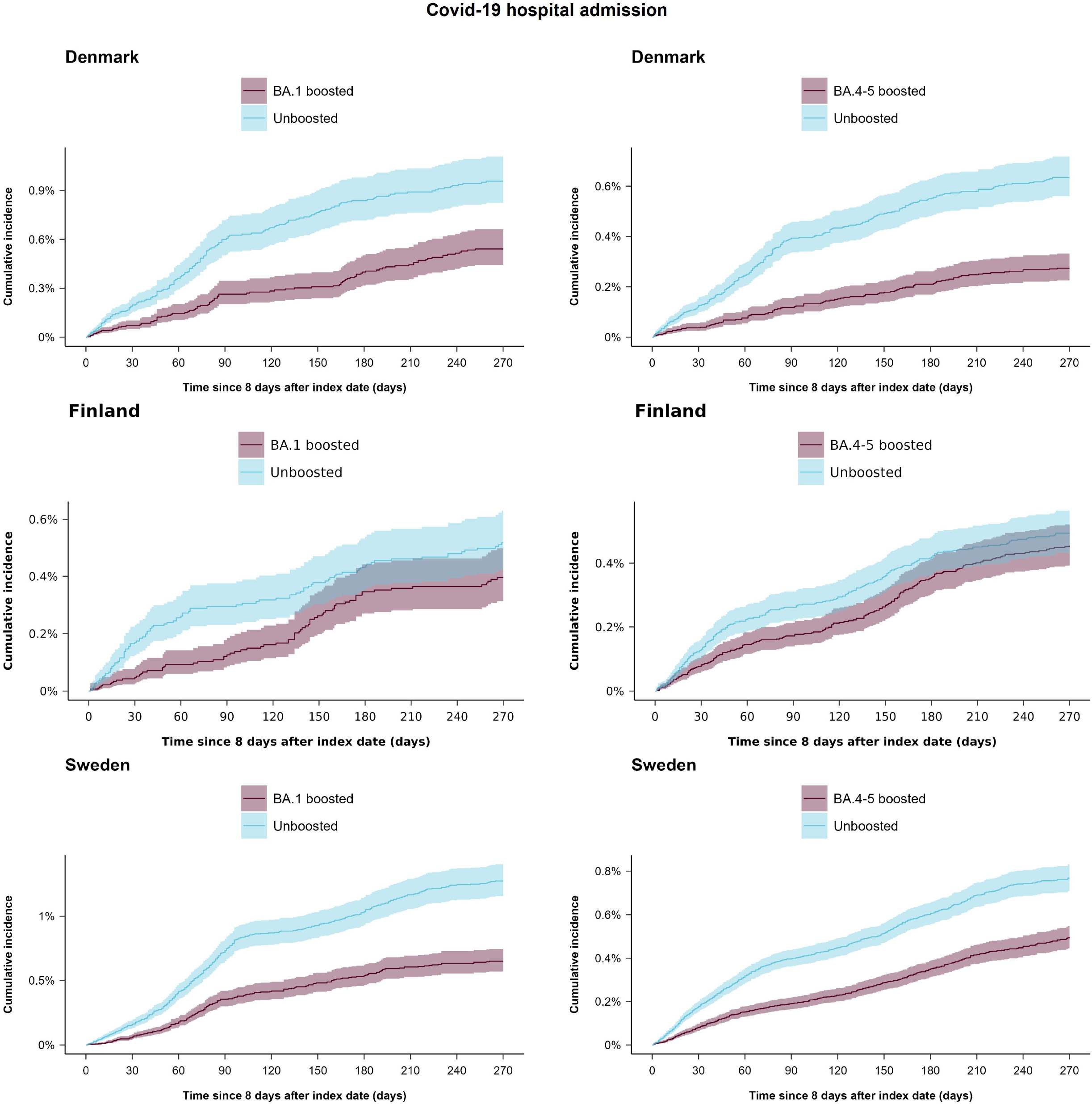

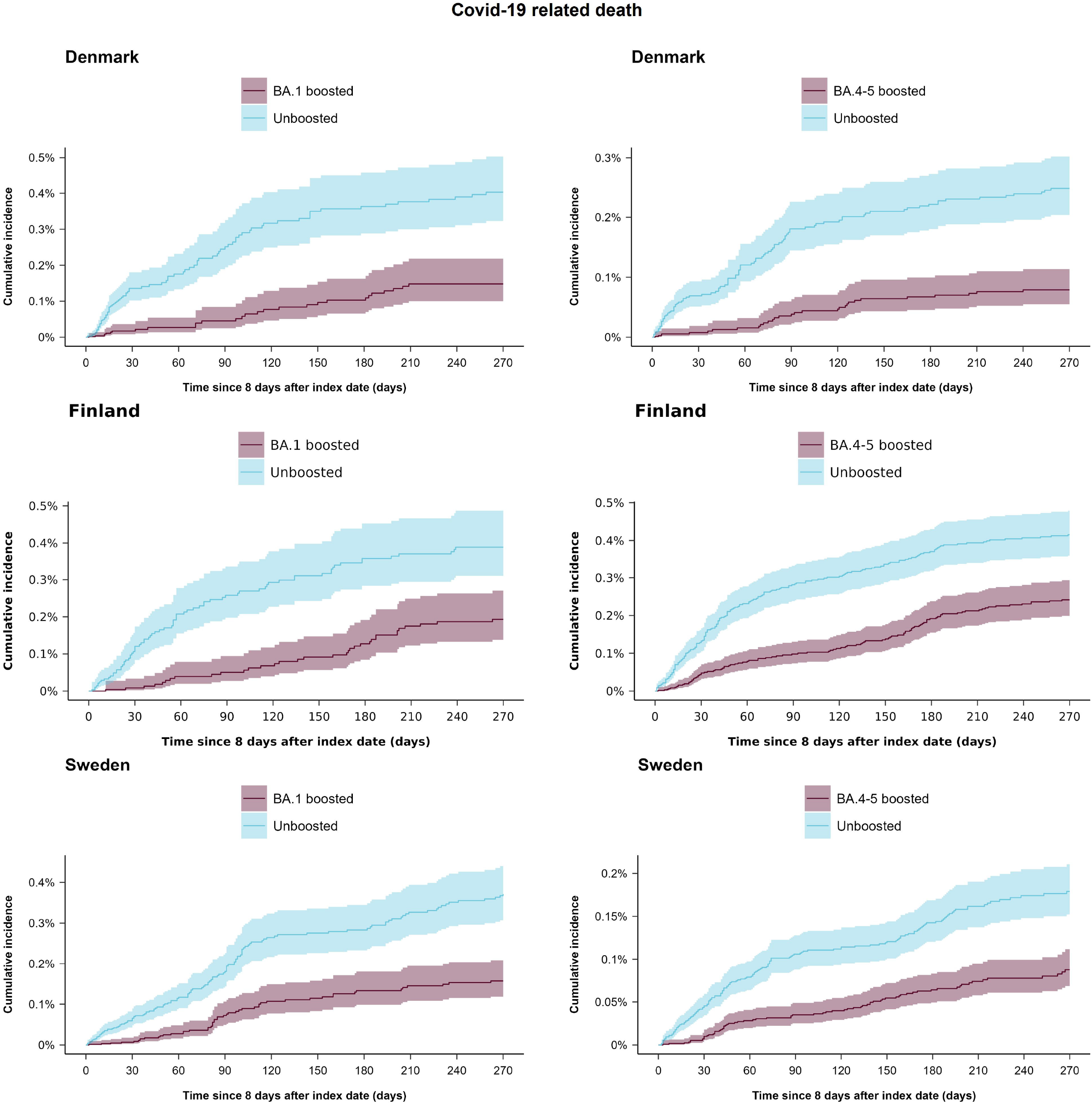

Table 2 presents the RD per 100,000 individuals and comparative VE against Covid-19 hospitalization at day 270 of follow-up for the bivalent boosters. The comparative VE against Covid-19 hospitalization was 34.2% (95% CI, 7.1% to 61.3%) for the bivalent BA.4-5 vaccine (696 vs 1,128 events) and 42.6% (95% CI, 31.3% to 53.9%) for the BA.1 vaccine (395 vs 740 events) compared with matched unboosted individuals. The corresponding RDs were -223.7 (95% CI, -411.5 to -36.0) and -385.0 (95% CI, -673.4 to -96.6) per 100,000 individuals, respectively. The comparative VE was similar across sex (female or male), age (<70 years or ≥70 years), dose number at which the bivalent booster was received (fourth dose, fifth dose, or ≥sixth dose), and immunocompromised condition subgroups.

**Table 2.** Risk of Covid-19-related hospitalization at day 270 comparing bivalent BA.4-5 or BA.1 boosted as a ≥fourth vaccine dose compared with unboosted immunocompromised individuals in three Nordic countries.

|  | Contributing countries | Events/person-years |  | Risk difference (95% CI) per 100,000 individuals | Comparative vaccine effectiveness (95% CI) |
| --- | --- | --- | --- | --- | --- |
|  |  | ≥Fourth dose bivalent boosted | Unboosted |  |  |
| Bivalent BA.4-5 booster |  |  |  |  |  |
| All | DK, FI, SE | 696 / 112,620 | 1128 / 110,408 | -223.7 (-411.5 to -36.0) | 34.2 (7.1 to 61.3) |
| Subgroups |  |  |  |  |  |
| Female | DK, FI, SE | 272 / 60,359 | 463 / 58,765 | -199.5 (-399.3 to 0.3) | 42.6 (14.3 to 71.0) |
| Male | DK, FI, SE | 424 / 52,261 | 665 / 51,643 | -250.9 (-459.9 to -41.8) | 30.7 (2.4 to 58.9) |
| Age <70 yrs | DK, FI, SE | 160 / 52,198 | 258 / 51,758 | -92.7 (-302.0 to 116.6) | 42.0 (15.2 to 68.7) |
| Age ≥70 yrs | DK, FI, SE | 536 / 60,422 | 870 / 58,650 | -323.2 (-542.7 to -103.7) | 33.2 (7.9 to 58.6) |
| Bivalent booster vaccine dose |  |  |  |  |  |
| Fourth dose | DK, FI, SE | 141 / 43,984 | 275 / 43,202 | -144.2 (-373.7 to 85.2) | 46.2 (19.7 to 72.8) |
| Fifth dose | DK, FI, SE | 341 / 46,006 | 536 / 44,877 | -301.9 (-538.7 to -65.0) | 35.6 (10.1 to 61.2) |
| ≥Sixth dose | DK, FI, SE | 206 / 21,814 | 303 / 21,528 | -32.3 (-386.0 to 321.3) | 5.0 (-32.7 to 42.7) |
| Immunocompromised condition |  |  |  |  |  |
| Solid malignancy | DK, FI, SE | 237 / 48,830 | 401 / 48,302 | -172.6 (-320.3 to -24.9) | 37.3 (14.5 to 60.0) |
| Hematologic malignancy | DK, FI, SE | 96 / 8,025 | 139 / 7,957 | -321.0 (-614.6 to -27.4) | 28.6 (1.3 to 55.9) |
| Rheumatologic or inflammatory disorder | DK, FI, SE | 142 / 30,498 | 246 / 29,383 | -193.3 (-352.1 to -34.6) | 41.8 (18.2 to 65.4) |
| Other intrinsic immune condition or immunodeficiency | FI, SE | 28 / 5,472 | 52 / 5,230 | -121.6 (-416.0 to 172.7) | 20.7 (-27.6 to 69.1) |
| Organ or stem cell transplant | DK, FI, SE | 36 / 2,352 | 36 / 2,276 | 47.8 (-446.2 to 541.9) | 11.6 (-37.5 to 60.6) |
| ≥2 of abovementioned conditions | DK, FI, SE | 77 / 5,668 | 142 / 5,428 | -748.8 (-1,118.7 to -378.9) | 50.1 (25.4 to 74.8) |
| Received a Covid-19 vaccine dose equivalent of booster dose for immunocompromised | DK, FI, SE | 80 / 11,775 | 112 / 11,832 | -187.0 (-391.1 to 17.2) | 27.4 (-3.0 to 57.9) |
| Bivalent BA.1 booster |  |  |  |  |  |
| All | DK, FI, SE | 395 / 49,801 | 740 / 48,172 | -385.0 (-673.4 to -96.6) | 42.6 (31.3 to 53.9) |
| Subgroups |  |  |  |  |  |
| Female | DK, FI, SE | 161 / 27,237 | 314 / 25,773 | -326.2 (-618.6 to -33.9) | 48.2 (36.1 to 60.4) |
| Male | DK, FI, SE | 234 / 22,564 | 426 / 22,400 | -441.7 (-751.8 to -131.6) | 39.2 (27.2 to 51.2) |
| Age <70 yrs | DK, FI, SE | 109 / 22,755 | 180 / 22,645 | -138.4 (-427.7 to 150.9) | 37.8 (20.5 to 55.2) |
| Age ≥70 yrs | DK, FI, SE | 286 / 27,046 | 560 / 25,528 | -548.6 (-853.4 to -243.8) | 44.7 (32.7 to 56.7) |
| Bivalent booster vaccine dose |  |  |  |  |  |
| Fourth dose | DK, FI, SE | 132 / 24,638 | 254 / 24,003 | -224.9 (-509.7 to 59.8) | 44.4 (30.8 to 58.0) |
| Fifth dose | DK, FI, SE | 238 / 23,737 | 437 / 22,797 | -490.9 (-793.1 to -188.7) | 44.3 (32.8 to 55.9) |
| ≥Sixth dose | FI, SE | 24 / 1,407 | 48 / 1,353 | -461.7 (-1,201.9 to 278.4) | 59.9 (36.5 to 83.2) |
| Immunocompromised condition |  |  |  |  |  |
| Solid malignancy | DK, FI, SE | 117 / 22,200 | 262 / 21,140 | -378.6 (-627.2 to -130.0) | 55.9 (45.6 to 66.2) |
| Hematologic malignancy | DK, FI, SE | 65 / 3,619 | 97 / 3,725 | -452.5 (-968.0 to 63.0) | 32.4 (9.7 to 55.0) |

|  | Contributing countries | Events/person-years |  | Risk difference (95% CI) per 100,000 individuals | Comparative vaccine effectiveness (95% CI) |
| --- | --- | --- | --- | --- | --- |
|  |  | ≥Fourth dose bivalent boosted | Unboosted |  |  |
| Rheumatologic or inflammatory disorder | DK, FI, SE | 93 / 13,601 | 157 / 12,848 | -256.3 (-527.1 to 14.6) | 46.0 (31.3 to 60.7) |
| Other intrinsic immune condition or immunodeficiency | DK, FI, SE | 21 / 2,460 | 41 / 2,359 | -382.2 (-833.6 to 69.2) | 43.3 (12.7 to 73.9) |
| Organ or stem cell transplant | DK, FI, SE | 26 / 922 | 29 / 1,013 | 130.4 (-822.8 to 1,083.7) | 49.7 (10.2 to 89.1) |
| ≥2 of abovementioned conditions | DK, FI, SE | 38 / 2,435 | 105 / 2,536 | -1,115.0 (-1,727.5 to -502.6) | 60.7 (45.2 to 76.1) |
| Received a Covid-19 vaccine dose equivalent of booster dose for immunocompromised | DK, FI, SE | 35 / 4,564 | 49 / 4,551 | -218.5 (-564.4 to 127.4) | 27.3 (-6.4 to 61.1) |

DK denotes Denmark, FI Finland, and SE Sweden.

Table 3 presents the RD per 100,000 individuals and comparative VE against Covid-19-related death at day 270 comparing bivalent boosted with unboosted immunocompromised individuals. The comparative VE against Covid-19-related death was 53.9% (95% CI, 38.6% to 69.3%) for the bivalent BA.4-5 vaccine (203 vs 457 events) and 57.9% (95% CI, 48.5% to 67.4%) for the BA.1 vaccine (112 vs 302 events) compared with matched unboosted. The RD was -138.7 (95% CI, -195.5 to -81.9) and -220.6 (95% CI, -275.9 to -165.4) per 100,000 individuals, respectively. The comparative VE was similar across sex (female or male), age (<70 years or ≥70 years), dose number at which the bivalent booster was received (fourth dose, fifth dose, or ≥sixth dose), and immunocompromised condition subgroups.

**Table 3.** Risk of Covid-19-related death at day 270 comparing bivalent BA.4-5 or BA.1 boosted as a ≥fourth vaccine dose compared with unboosted immunocompromised individuals in three Nordic countries.

| Contributing countries | Events/person-years | Risk difference (95% CI) | Comparative |
| --- | --- | --- | --- |

|  | ting<br>countries | ≥Fourth dose<br>bivalent<br>boosted | Unboosted | per 100,000 individuals | vaccine<br>effectiveness (95%<br>CI) |
| --- | --- | --- | --- | --- | --- |
| **Bivalent BA.4-5 booster** |  |  |  |  |  |
| All | DK, FI, SE | 203 / 113,540 | 457 / 111,383 | -138.7 (-195.5 to -81.9) | 53.9 (38.6 to 69.3) |
| Subgroups |  |  |  |  |  |
| Female | DK, FI, SE | 83 / 60,913 | 176 / 59,233 | -104.8 (-162.4 to -47.1) | 54.2 (36.3 to 72.1) |
| Male | DK, FI, SE | 120 / 52,627 | 281 / 52,150 | -174.2 (-241.5 to -107.0) | 53.7 (37.2 to 70.2) |
| Age <70 yrs | DK, FI, SE | 30 / 52,167 | 67 / 51,797 | -48.2 (-98.6 to 2.2) | 59.3 (36.9 to 81.7) |
| Age ≥70 yrs | DK, FI, SE | 173 / 61,373 | 390 / 59,586 | -195.1 (-261.9 to -128.4) | 52.5 (37.1 to 68.0) |
| Bivalent booster vaccine<br>dose |  |  |  |  |  |
| Fourth dose | DK, FI, SE | 57 / 44,238 | 125 / 43,468 | -73.7 (-153.5 to 6.1) | 47.5 (22.2 to 72.8) |
| Fifth dose | DK, FI, SE | 113 / 46,596 | 261 / 45,502 | -207.1 (-293.0 to -121.2) | 62.7 (41.3 to 84.2) |
| ≥Sixth dose | FI, SE | 33 / 21,996 | 71 / 21,719 | -161.8 (-328.9 to 5.2) | 50.6 (6.6 to 94.6) |
| Immunocompromised<br>condition |  |  |  |  |  |
| Solid malignancy | DK, FI, SE | 82 / 49,097 | 182 / 48,709 | -114.7 (-161.3 to -68.1) | 55.5 (38.2 to 72.9) |
| Hematologic malignancy | DK, FI, SE | 22 / 8,186 | 56 / 8,078 | -252.7 (-389.5 to -115.9) | 73.4 (53.5 to 93.4) |
| Rheumatologic or<br>inflammatory disorder | DK, FI, SE | 25 / 30,715 | 70 / 29,542 | -90.7 (-140.7 to -40.6) | 66.2 (43.7 to 88.6) |
| Other intrinsic immune<br>condition or<br>immunodeficiency | DK, FI, SE | 14 / 5,527 | 32 / 5,380 | -121.1 (-255.9 to 13.7) | 59.2 (30.4 to 88.1) |
| Organ or stem cell<br>transplant | DK, FI, SE | 7 / 2,444 | 8 / 2,330 | -48.4 (-264.6 to 167.9) | 23.8 (-60.4 to 100.0) |
| ≥2 of abovementioned<br>conditions | DK, FI, SE | 18 / 5,838 | 45 / 5,515 | -226.6 (-412.1 to -41.1) | 72.9 (48.1 to 97.7) |
| Received a Covid-19 vaccine<br>dose equivalent of booster<br>dose for<br>immunocompromised | DK, FI, SE | 35 / 11,733 | 64 / 11,828 | -64.2 (-141.4 to 13.0) | 45.3 (16.0 to 74.6) |
| **Bivalent BA.1 booster** |  |  |  |  |  |
| All | DK, FI, SE | 112 / 50,237 | 302 / 48,688 | -220.6 (-275.9 to -165.4) | 57.9 (48.5 to 67.4) |
| Subgroups |  |  |  |  |  |
| Female | DK, FI, SE | 48 / 27,251 | 119 / 26,049 | -149.8 (-216.5 to -83.0) | 57.9 (42.7 to 73.2) |
| Male | DK, FI, SE | 64 / 22,986 | 183 / 22,639 | -297.0 (-387.7 to -206.3) | 60.4 (48.6 to 72.1) |
| Age <70 yrs | DK, FI, SE | 18 / 22,812 | 38 / 22,745 | -40.1 (-82.6 to 2.3) | 48.5 (18.4 to 78.5) |
| Age ≥70 yrs | DK, FI, SE | 94 / 27,425 | 264 / 25,943 | -358.2 (-454.4 to -262.0) | 59.7 (49.7 to 69.8) |
| Bivalent booster vaccine<br>dose |  |  |  |  |  |
| Fourth dose | DK, FI, SE | 43 / 24,786 | 101 / 24,183 | -90.8 (-227.4 to 45.7) | 63.3 (47.0 to 79.6) |
| Fifth dose | DK, FI, SE | 64 / 23,997 | 186 / 23,101 | -323.3 (-475.5 to -171.1) | 61.7 (49.1 to 74.4) |
| ≥Sixth dose | FI, SE | 5 / 1,454 | 15 / 1,404 | -425.7 (-866.8 to 15.3) | 64.1 (26.1 to 100.0) |
| Immunocompromised<br>condition |  |  |  |  |  |
| Solid malignancy | DK, FI, SE | 38 / 22,388 | 132 / 21,369 | -243.6 (-322.9 to -164.4) | 65.9 (53.1 to 78.6) |
| Hematologic malignancy | DK, FI, SE | 21 / 3,709 | 36 / 3,809 | -148.0 (-419.7 to 123.6) | 44.9 (11.2 to 78.7) |
| Rheumatologic or<br>inflammatory disorder | DK, FI, SE | 22 / 13,667 | 47 / 12,884 | -128.4 (-213.8 to -43.1) | 56.4 (32.8 to 80.0) |
| Other intrinsic immune<br>condition or | DK, FI | 5 / 2,447 | 23 / 2,437 | -471.7 (-928.8 to -14.6) | 87.6 (68.3 to 100.0) |

**Table 3. Risk of Covid-19-related death at day 270 comparing bivalent BA.4-5 or BA.1 boosted as a ≥ fourth vaccine dose compared with unboosted immunocompromised individuals in three Nordic countries.**
|  | Contributing countries | Events/person-years |  | Risk difference (95% CI) per 100,000 individuals | Comparative vaccine effectiveness (95% CI) |
| --- | --- | --- | --- | --- | --- |
| | | ≥Fourth dose bivalent boosted | Unboosted | | |
| immunodeficiency |  |  |  |  |  |
| Organ or stem cell transplant | DK, FI, SE | <5 / 944 | <5 / 1,049 | NE | NE |
| $\geq 2$ of abovementioned conditions | DK, FI, SE | 14 / 2,464 | 32 / 2,520 | -221.9 (-520.6 to 76.7) | 51.3 (15.9 to 86.8) |
| Received a Covid-19 vaccine dose equivalent of booster dose for immunocompromised | DK, FI, SE | 10 / 4,617 | 30 / 4,619 | -226.9 (-433.6 to -20.3) | 53.6 (19.3 to 87.9) |

DK denotes Denmark, FI Finland, NE not estimable, and SE Sweden.

### Waning of bivalent booster vaccine effectiveness

Figure 3 shows the waning of the VE against Covid-19-related hospitalization and death. The comparative VE estimates were highest in the first 45 days and waned gradually during the 270 days of follow-up. The VE against Covid-19 hospitalization at the end of the first 45 days of follow-up was 52.8% (95% CI, 38.6% to 67.0%) for BA.4-5 boosted and 62.2% (95% CI, 53.5% to 70.9%) for BA.1 boosted compared with unboosted. The meta-regression indicated waning of the comparative VE of -9.3 (95% CI, -15.0 to -3.5) percentage points for BA.4-5 boosted and -5.5 (95% CI, -10.3 to -0.7) percentage points for BA.1 boosted per 45 days since eight days after bivalent booster vaccination. The VE against Covid-19-related death at the end of the first 45 days of follow-up was 72.8% (95% CI, 60.0% to 85.6%) for BA.4-5 boosted and 84.2% (95% CI, 76.6% to 91.7%) for BA.1 boosted compared with unboosted. The meta-regression indicated waning of the VE of - 9.2 (95% CI, -16.7 to -1.7) percentage points for BA.4-5 boosted and -6.2 (95% CI, -14.1 to 1.8) percentage points for BA.1 boosted per 45 days since eight days after bivalent booster vaccination.

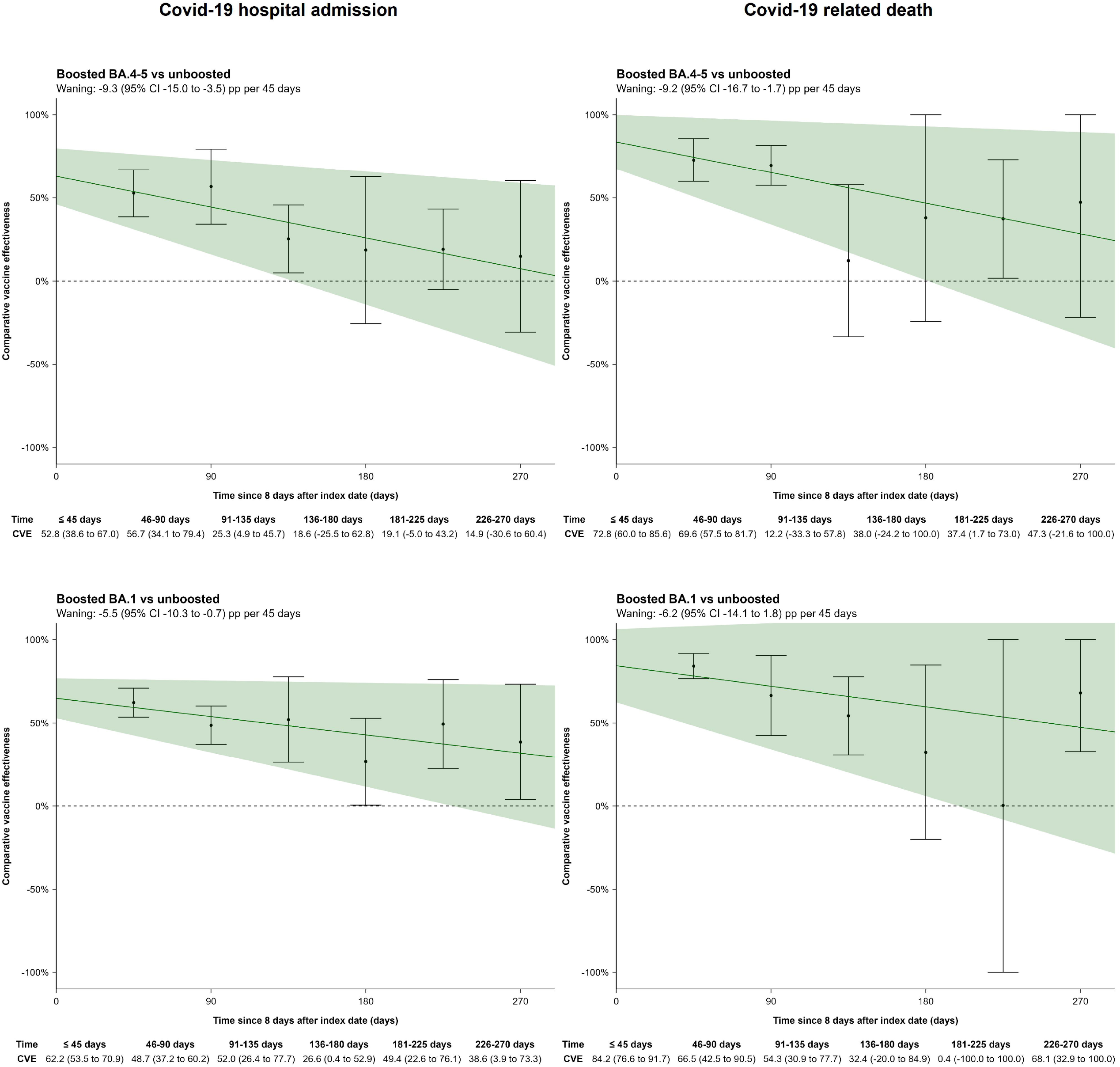

## Discussion

This nationwide cohort study estimated the comparative effectiveness of the bivalent BA.4-5 or BA.1 boosters against severe Covid-19 among immunocompromised across Denmark, Finland and Sweden. We found that immunocompromised individuals receiving a bivalent BA.4-5 or a BA.1 booster had lower risk of hospitalization and death related to Covid-19 during 9 months of follow-up compared with immunocompromised individuals who had not received the booster. We report overall moderate comparative VE estimates against Covid-19 hospitalization of 34% for bivalent BA.4-5 and 43% for bivalent BA.1 booster at end of the 9 months follow-up period, but slightly higher comparative VE estimates of 54% and 58% against Covid-19-related death, respectively. In addition, we observed that the comparative VE was highest during the first 45 days since vaccination (≥53% and ≥73% for Covid-19 hospitalization and death, respectively) with subsequent gradual waning. In contrast, the number of severe outcomes prevented among the boosted individuals were significant. At day 270 of follow-up, BA.4-5 and BA.1 boosters had prevented 223.7 and 385.0 Covid-19 hospitalizations and 138.7 and 220.6 Covid-19 deaths per 100,000 individuals. As such, these estimates correspond to number needed to vaccinate (NNV) to prevent one Covid-19-related hospitalization of 447 and 260 for bivalent BA.4-5 and BA.1 booster, respectively. The NNV to prevent one Covid-19-related death was 721 and 453 for bivalent BA.4-5 and BA.1 booster, respectively.

Few studies have evaluated the effectiveness of the bivalent boosters among immunocompromised populations (5, 6, 12). Similar to our findings, a matched cohort study in immunocompromised individuals conducted in the US observed 65% (95% CI, 44% to 78%) vaccine effectiveness (VE) for the bivalent BA.4-5 vaccine against Covid-19 hospitalization compared with ≥2 doses of Original monovalent mRNA vaccine (12). A US test-negative case-control study of individuals aged ≥18 years with immunocompromising conditions who were hospitalized with Covid-19-like illness reported a VE against laboratory-confirmed Covid-19–associated hospitalization of 28% (95% CI, 10% to 42%) during the 7-59 days after bivalent BA.4-5 booster compared with unvaccinated (5). The VE decreased to 13% (95% CI, -13% to 33%) during the 120-179 days (5). Moreover, another US study of immunocompromised patients reported a VE comparing bivalent BA.4-5 booster vaccinated to unvaccinated of 78% (95% CI, 20% to 94%) against Covid-19 hospitalization with the subvariant BA.4-5 (6). In the same study, the VE was 69% (95% CI, -15% to 92%) against Covid-19 hospitalization with the subvariant XBB (6). The interpretation of both of these US studies are complicated by the comparison to unvaccinated (i.e., no history of any prior Covid-19 vaccine) immunocompromised individuals and the high likelihood of selection bias.

Our results contribute to the existing literature by providing data on long-term relative and absolute effectiveness, waning effectiveness, and effectiveness across subgroups of relevance to national vaccination policy. Despite the moderate VE at day 270, the absolute benefits of vaccination, as reflected by the risk differences at day 270, was significant due to the higher baseline risk of poor Covid-19 outcome in this population of immunocompromised.

### Strengths and limitations

This study benefits from the completeness of the nationwide registries, ensuring correct individual-level data linkage between registries. We used a matched study design and compared bivalent booster vaccinated with unboosted that had previously received the same number of Covid-19 vaccines to mitigate confounding, thereby strengthening the internal validity of the results. This method is likely superior to previous studies comparing bivalent boosted to unvaccinated individuals as unvaccinated immunocompromised individuals at this stage in the pandemic are likely to be highly selected and not comparable to the immunocompromised population getting vaccinated. Still, the possibility of residual and unmeasured confounding factors cannot be excluded. Also, there is a potential for healthy vaccinee bias, wherein individuals who choose to get vaccinated may inherently possess healthier behaviors or lifestyles, leading to an overestimation of the true VE and RD. Healthy vaccine bias may also manifest when vaccination is postponed for people who are seriously ill. In an immunocompromised population, we will also need to take into account the fact that those with more severe manifestations of immunocompromising conditions may be more likely to get vaccinated due to a higher perceived personal risk. This will tend to underestimate the true VE and RD.

While both outcomes under study relied on PCR testing, differences in testing strategies across countries or hospitals may lead to variations in the detection of Covid-19 hospitalization and Covid-19-related death. This variability could result in underreporting of events. Furthermore, the outcome definitions might have included individuals whose condition was not directly linked to Covid-19, but where Covid-19 either contributed to the outcome or coincided with hospital admission or death. The potential outcome misclassification may affect the VE estimates (13). Similarly, we could not take hybrid immunity into account as not all SARS-CoV-2 infections are documented by PCR tests due to the testing strategy, especially after the initial big Omicron waves and the subsequent relaxation of testing indications in the Nordic countries (14). We expect that any potential proportion of unmeasured recent history SARS-CoV-2 infection would be higher among the unboosted relative to the boosted group, which would tend to bias our results toward lower effectiveness. Potential outcome misclassifications are expected to be non-differential between the actively compared groups, and would as such, skew towards conservative estimates.

The identified immunocompromised individuals for our study cohorts were presumed immunocompromised, which we ascertained by disease diagnoses or Covid-19 vaccination patterns. However, immunocompromised populations are heterogeneous and can involve both diseases and treatments (e.g. treatment for autoimmune conditions or cancer chemotherapy). Moreover, the exact definitions differed between countries. Additionally, some individuals might only be temporary or short-term immunocompromised due to the duration of immunosuppressive treatment or the recency of transplant surgery. Thereby, the degree of immunosuppression likely varied within this cohort and it also might have included individuals not truly immunocompromised. In addition, it might well be that those with more severe immunocompromising conditions are more likely to get vaccinated, which would bias towards lower observed effectiveness, since these patients have a higher baseline risk of severe outcomes. Our results likely have a high degree of generalizability to similarly defined of immunocompromised with the same age distribution as our immunocompromised cohort (a high mean age of 71-72 years). While the effectiveness appeared similar across subgroups examined, some estimates were imprecise due to few events. The mean age in our cohort was high compared to the inclusion criteria of 18 years or above. Furthermore, the results may differ in periods with other SARS-CoV-2 variants.

## Conclusions

Among adults with immunocompromised conditions in Denmark, Finland, and Sweden, vaccination with a bivalent BA.4-5 or BA.1 booster lowered the risk of Covid-19-related hospitalization and death over a follow-up period of 9 months. The effectiveness did not differ across subgroups and was highest during the first months since vaccination with subsequent gradual waning.

## Data Availability

Owing to data privacy regulations in each country, the raw data cannot be shared.

## Contributors

NWA, EMT, and AH conceptualized the study. MAG drafted the manuscript. EMT, NP, and JP did the statistical analysis. All authors interpreted the results and critically reviewed the manuscript. AH supervised the study. MAG, NWA, EMT, and AH are the guarantors.

## Acknowledgement

We thank Ulrike Baum and Toni Lehtonen for assisting in the analyses and in the data collection.

## Ethics approval

Denmark: The Danish analyses were performed as surveillance activities analyses as part of the advisory tasks of the governmental institution Statens Serum Institut (SSI) for the Danish Ministry of Health. SSI’s purpose is to monitor and fight the spread of disease in accordance with section 222 of the Danish Health Act. According to Danish law, national surveillance activities conducted by SSI do not require approval from an ethics committee. Both the Danish Governmental law firm and the compliance department of SSI have approved that the study is fully compliant with all legal, ethical, and IT-security requirements and there are no further approval procedures required for such studies.

Finland: By Finnish law, the Finnish Institute for Health and Welfare (THL) is the national expert institution to carry out surveillance of the impact of vaccinations in Finland (Communicable Diseases Act, https://www.finlex.fi/en/laki/kaannokset/2016/en20161227.pdf). Neither specific ethical approval of this study nor informed consent from the participants were needed.

Sweden: The Swedish analyses were conducted under the Swedish Ethical Review Authority approval 2020-06859, 2021-02186 and conformed to the principles embodied in the Declaration of Helsinki. Register-based studies (like this) in Sweden are exempt from obtaining consent to participate.

## Competing interests

EP reports receiving a unrelated grant from Finnish Medical Foundation.

RL reports receiving grants from Sanofi Aventis paid to his institution outside the submitted work; and receiving personal fees from Pfizer outside the submitted work.

NP and RL are employed at the Swedish Medical Products Agency, Uppsala, Sweden. The views expressed in this paper do not necessarily represent the views of the Government agency.

AH reports receiving unrelated grants from Independent Research Fund Denmark, the Lundbeck Foundation and the Novo Nordisk Foundation. AH reports acting as a scientific board member for VAC4EU.

### Data sharing

Owing to data privacy regulations in each country, the raw data cannot be shared. Transparency

The lead author (the manuscript’s guarantor) affirms that this manuscript is an honest, accurate, and transparent account of the study being reported; that no important aspects of the study have been omitted; and that any discrepancies from the study as planned (and, if relevant, registered) have been explained.

## References

1. Andersson NW, Thiesson EM, Baum U, Pihlström N, Starrfelt J, Faksová K, et al. Comparative effectiveness of bivalent BA.4-5 and BA.1 mRNA booster vaccines among adults aged ≥50 years in Nordic countries: nationwide cohort study. BMJ. 2023;382:e075286.

2. Tartof SY, Slezak JM, Puzniak L, Hong V, Frankland TB, Ackerson BK, et al. Effectiveness of BNT162b2 BA.4/5 bivalent mRNA vaccine against a range of COVID-19 outcomes in a large health system in the USA: a test-negative case-control study. Lancet Respir Med. 2023;11(12):1089–100.

3. Lin DY, Xu Y, Gu Y, Zeng D, Wheeler B, Young H, et al. Effectiveness of Bivalent Boosters against Severe Omicron Infection. N Engl J Med. 2023;388(8):764–6.

4. Arbel R, Peretz A, Sergienko R, Friger M, Beckenstein T, Duskin-Bitan H, et al. Effectiveness of a bivalent mRNA vaccine booster dose to prevent severe COVID-19 outcomes: a retrospective cohort study. Lancet Infect Dis. 2023;23(8):914–21.

5. Link-Gelles R, Weber ZA, Reese SE, Payne AB, Gaglani M, Adams K, et al. Estimates of Bivalent mRNA Vaccine Durability in Preventing COVID-19–Associated Hospitalization and Critical Illness Among Adults with and Without Immunocompromising Conditions — VISION Network, September 2022– April 2023. American Journal of Transplantation. 2023;23(7):1062–76.

6. Ackerson BK, Bruxvoort KJ, Qian L, Sy LS, Qiu S, Tubert JE, et al. Effectiveness and durability of mRNA-1273 BA.4/BA.5 bivalent vaccine (mRNA-1273.222) against SARS-CoV-2 BA.4/BA.5 and XBB sublineages. medRxiv. 2023:2023.12.11.23299663.

7. Szekanecz Z, Vokó Z, Surján O, Rákóczi É, Szamosi S, Szűcs G, et al. Effectiveness and waning of protection with the BNT162b2 vaccine against the SARS-CoV-2 Delta variant in immunocompromised individuals. Front Immunol. 2023;14:1247129.

8. Di Fusco M, Lin J, Vaghela S, Lingohr-Smith M, Nguyen JL, Scassellati Sforzolini T, et al. COVID-19 vaccine effectiveness among immunocompromised populations: a targeted literature review of real-world studies. Expert Rev Vaccines. 2022;21(4):435–51.

9. Zheng Z, Peng F, Xu B, Zhao J, Liu H, Peng J, et al. Risk factors of critical & mortal COVID-19 cases: A systematic literature review and meta-analysis. Journal of Infection. 2020;81(2):e16–e25.

10. Martono, Fatmawati F, Mulyanti S. Risk Factors Associated with the Severity of COVID-19. Malays J Med Sci. 2023;30(3):84–92.

11. Sera F, Armstrong B, Blangiardo M, Gasparrini A. An extended mixed-effects framework for meta-analysis. Stat Med. 2019;38(29):5429–44.

12. Tseng HF, Ackerson BK, Sy LS, Tubert JE, Luo Y, Qiu S, et al. mRNA-1273 bivalent (original and Omicron) COVID-19 vaccine effectiveness against COVID-19 outcomes in the United States. Nat Commun. 2023;14(1):5851.

13. Hansen CH. Bias in vaccine effectiveness studies of clinically severe outcomes that are measured with low specificity: the example of COVID-19-related hospitalisation. Euro Surveill. 2024;29(7).

14. Gram MA, Steenhard N, Cohen AS, Vangsted AM, Mølbak K, Jensen TG, et al. Patterns of testing in the extensive Danish national SARS-CoV-2 test set-up. PLoS One. 2023;18(7):e0281972.

