## Supplementary for "Comparative effectiveness of bivalent BA.4.5 or BA.1 mRNA booster vaccines among immunocompromised individuals across three Nordic countries: a nationwide cohort study"

**Supplementary Appendix**

### Supplementary Table S1. Definition of immunocompromised conditions.

| **Immunocompromised condition** |  | **Definition** |
| --- | --- | --- |
| Solid malignancy | Denmark, Sweden | ICD-10 codes: C00–C80 (except C44) registered within 3 years prior to start of the study period |
|  | Finland | ICD-10 codes: C00–C43, C45–C80, C97, D05.1, D39 registered within 2 years prior to the start of the study period |
| Hematologic malignancy | Denmark, Sweden | ICD-10 codes: C81–C86, C88, C90–C96, D46, D47, D61.0, D70.0, D61.2, D61.9, D71 registered within 3 years prior to the start of the study period |
|  | Finland | ICD-10 codes: C81–C85, C88, C90–C96 registered within 2 years prior to the start of the study period. |
| Rheumatologic or inflammatory disorder | Denmark, Sweden | ICD-10 codes: D86, E85 [except E85.0], G35, J67.9, L40.1, L40.5, L93, L94, M05–M08, M30, M31.3, M31.5, M32–M35, M46 registered within 3 years prior to the start of the study period |
|  | Finland | ICD-10 codes: D86, K50, K51, L40, M02, M05–M07, M13.9, M45, M46.0, M46.1, M46.9, M94.1 registered prior to the start of the study period or prescription of H02AB02, H02AB04, H02AB06, H02AB07, L04AA06, L04AA10, L04AA13, L04AA18, L04AA24, L04AA26, L04AA29, L04AA33, L04AA37, L04AB, L04AC, L04AD01, L04AD02, L04AX01, L04AX03, L04AB, L01XC02, L04AA24, L04AA26, L04AA33, L04AC before 27 December 2020. |
| Other intrinsic immune condition or immunodeficiency | Denmark, Sweden | ICD-10 codes: B9735, D27.9, D61, D72.8, D80, D81 [except D81.3], D82–D84, D89 [except D89.2], K70.3, K70.4, K72, K74.3–K74.6 [except K74.60 and K74.69], N04, R18, Z992 registered within 3 years prior to start of the study period |
|  | Finland | ICD-10 codes: K70.2, K70.3, K70.4, K71–K74, D70.8, D80–D84, E31.00 registered prior to the start of the study period |
| Organ or stem cell transplant | Denmark, Sweden | ICD-10 codes: T86 [except T86.82–T86.84, T86.89, and T86.9], Z94, and Z98.85 registered within 3 years prior start of the study period |
|  | Finland | ICD-10 codes: T86, Z94 registered prior to the start of the study period. |
| Received a Covid-19 vaccine dose equivalent of booster dose for immunocompromised | Denmark, Finland, Sweden | Either 1) any booster dose (≥3rd dose) within 90 days of the last dose^a^, 2) receipt of fourth dose before the roll-out of the 4th dose boosters^b^, or 3) receipt of fifth or more vaccine dose doses prior to start of study period |

Definitions of immunocompromised condition 1 to 5 were adapted and modified from Embi et al. (1) and Hughes et al. (2) and Salo et al. in Finland (3). ^a^Covid-19 vaccination courses in the Nordic countries for the general population have generally had time intervals of 6 months or more between boosters. ^b^Of note, the start date for the roll-out of the fourth dose was earlier than start of the study period in Finland and Sweden, as vulnerable and the elderly living in nursing homes were initially prioritized during Spring 2022, before the fourth dose was rolled out to the general population. The exact start date of the rollout was set to 31 March 2022 in Finland, and 15 February 2022 in Sweden, while coinciding with the start of the study period in Denmark (being 1 September 2022).

### Supplementary Table S2. Definition of outcomes.

| **Outcome variable** | **Country** | **Data source and details** |
| --- | --- | --- |
| Covid-19 hospitalization | Denmark | *The National Patient Register and the Danish Microbiology Database.* Defined as a hospitalization with a PCR positive test for SARS-CoV-2 within 14 days before to 2 days after the admission date, b) inpatient contact or at least 12 hours of contact, and c) a Covid-19 relevant diagnosis code (ICD-10: B342, B342A, B948A, B972, B972A, B972B, B972B1, Z038PA1) |
|  | Finland | *National Care Register for Health Care and the National Infectious Diseases Register.* Defined as a hospitalization with a PCR positive test for SARS-CoV-2 within 14 days before to 2 days after the admission date, b) inpatient hospital contact, and c) a Covid-19 relevant main diagnosis (ICD-10: J00-J22, J46, J80-J84, J851, J86, U071, U072). |
|  | Sweden | *The Swedish Patient Register and the Register on surveillance of notifiable communicable diseases (SmiNet).* Defined as a hospitalization with a PCR positive test for SARS-CoV-2 within 14 days before to 2 days after the admission date, b) inpatient contact or at least 12 hours of contact, and c) a Covid-19 relevant diagnosis code (ICD-10: U071, U072, U109) |
| Covid-19-related death | Denmark | *The Civil Registration System and the Danish Microbiology Database.* Defined as (the date of) death within 30 days after PCR positive test for SARS-CoV-2. |
|  | Finland | *The Finnish Population Information System and the National Infectious Diseases Register.* Defined as (the date of) death within 30 days after PCR positive test for SARS-CoV-2. |
|  | Sweden | *The Total Population Register, the Cause of Death Register, and the Swedish Patient Register and the Register on surveillance of notifiable communicable diseases (SmiNet).* Defined as (the date of) death within 30 days after PCR positive test for SARS-CoV-2. |

### Supplementary Table S3. Definition of covariates.

| **Variable** | **Country** | **Data source and details** | **Values/codes** |
| --- | --- | --- | --- |
| Age | Denmark | *The Civil Registration System.* Recorded birth year. Age defined as the country-specific start date minus birth year. | Categorical (for adjustment, using birth year): 5-year bins  Binary (for stratification): </≥ 70 years |
|  | Finland | *The Finnish Population Information System.* Recorded birth year. Age defined as the country-specific start date minus birth year. |  |
|  | Sweden | *The Total Population Register.* Recorded birth year. Age defined as the country-specific start date minus birth year. |  |
| Sex | Denmark | *The Civil Registration System.* Defined as registered sex. | Binary: male, female |
|  | Finland | *The Finnish Population Information System.* Defined as registered sex. |  |
|  | Sweden | *The Total Population Register.* Defined as registered sex. |  |
| Calendar month of last monovalent dose received | Denmark | *The Danish Vaccination Register.* Defined by the date where the respective vaccine dose examined was administered (i.e., third or fourth dose). | Categorical (monthly [up to 34 levels]): 1 (27 December 2020-31 January 2021) to month 34 (October 2023) |
|  | Finland | *The National Vaccination Register.* Defined by the date where the respective vaccine dose examined was administered (i.e., third or fourth dose). |  |
|  | Sweden | *The National Vaccination Register.* Defined by the date where the respective vaccine dose examined was administered i.e., third or fourth dose). |  |
| Region of residency | Denmark | *The Civil Registration System.* Defined by last known address at the country-specific start date for the rollout of the fourth vaccine dose. | Categorical: Denmark, 5 levels; Finland, 5 levels; Sweden, 9 levels |
|  | Finland | *The Finnish Population Information System.* Defined by last known municipality of residence. |  |
|  | Sweden | *The Total Population Register.* Defined by last known address at the country-specific start date for the rollout of the fourth vaccine dose. |  |
| Comorbidity 1: Chronic pulmonary disease | Denmark | *The National Patient Register.* Defined as primary diagnoses regardless of type of hospital contact registered before the start date for the country-specific rollout of the fourth vaccine dose (look-back 3 years). | Binary: yes/no (ICD-10 codes: J40-J47, J60–J67, J684, J701, J703, J841, J920, J961, J982, J983) |
|  | Finland | *Care register for Health Care.* Defined as primary or secondary diagnoses registered prior to the start of the study period. | Binary: yes/no (ICD-10 codes: J41-J44, J47) |
|  | Sweden | *National Patient Register.* Defined as any recorded ICD-10 diagnosis during inpatient or outpatient contact and before first Covid-19 vaccination (look-back 3 years). | Binary: yes/no (ICD-10 codes: E84, J41-J47, J84, J98) |
| Comorbidity 2: Cardiovascular conditions and diabetes | Denmark | *The National Patient Register.* Defined as primary diagnoses regardless of type of hospital contact registered before the start date for the country-specific rollout of the fourth vaccine dose (look-back 3 years). | Binary: yes/no (ICD-10 codes: E10-E11, I110, I130, I132, I20-I23, I420, I426-I429, I48, I500-I503, I508, I509) |
|  | Finland | *Care register for Health Care, Register of Primary Health Care Visits, Special Reimbursement Register and Prescription Centre database.* Defined as primary or secondary diagnoses prior to the start of the study period or drug prescriptions before 27 December 2020. | Binary: yes/no (ICD-10 codes: E10, E11, E13-E14, I11–I13, I15, I20–I25; ICPC-2 codes: T89, T90; ATC codes: A10A, A10B) |
|  | Sweden | *National Patient Register.* Defined as any recorded ICD-10 diagnosis during inpatient or outpatient contact and before first Covid-19 vaccination (look-back 3 years).  *Swedish Prescribed Drug Register.* Antidiabetic drugs use defined as ≥2 filled prescriptions during 2020. | Binary: yes/no (ICD-10 codes: E10-E14, I05-I09, I110, I20-I28, I34-I37, I39, I42, I43, I46, I48-I50; ATC code: A10) |
| Comorbidity 3: Selected autoimmunity-related conditions^a^ | Denmark | *The National Patient Register.* Defined as primary diagnoses regardless of type of hospital contact registered before the start date for the country-specific rollout of the fourth vaccine dose (look-back 3 years). | Binary: yes/no (ICD-10 codes: D510, D590, D591, D690, D693, D86, E050, E063, E271, E272, G122G, G35, G610, G700, I00, I01, K50, K51, K743, K900, L12, L40, L52, L80, L93, M05, M06, M08, M300, M313, M315, M316, M32, M33, M34, M35, M45) |
|  | Finland | *Care register for Health Care, Special Reimbursement Register and Prescription Centre database.* Defined as primary or secondary diagnoses prior start of the follow-up or drug prescriptions before 27 December 2020. | Binary: yes/no (ICD-10 codes: D7081, D7089, D80–D84, E250, E271, E272, E274, E310, E896, D86, K50, K51, L40, M02, M05–M07, M139, M45, M460, M461, M469, M941; ATC-codes: H02AB02, H02AB04, H02AB06, H02AB07, L01BA01, L01XC02, L04AA06, L04AA10, L04AA13, L04AA18, L04AA24, L04AA26, L04AA29, L04AA33, L04AA37, L04AB, L04AC, L04AD01, L04AD02, L04AX01, L04AX03) |
|  | Sweden | *National Patient Register.* Defined as any recorded ICD-10 diagnosis during inpatient or outpatient contact and before first Covid-19 vaccination (look-back 3 years). | Binary: yes/no (ICD-10 codes: D86, G35, K50, K51, L40, M05-M09, M13, M14, M45) |
| Comorbidity 4: Cancer | Denmark | *The National Patient Register.* Defined as primary diagnoses regardless of type of hospital contact registered before the start date for the country-specific rollout of the fourth vaccine dose (look-back 3 years). | Binary: yes/no (ICD-10 codes: C00–C85 (without C44), C88, C90-C96) |
|  | Finland | *Care register for Health Care and Special Reimbursement Register.* Defined as primary or secondary diagnoses registered within 2 years prior to the start of the study period. | Binary: yes/no (ICD-10 codes: C00–C43, C45–C80, C97, D05.1, D39) |
|  | Sweden | *National Patient Register.* Defined as any recorded ICD-10 diagnosis during inpatient or outpatient contact and before first Covid-19 vaccination (look-back 3 years). | Binary: yes/no (ICD-10 codes: C00-C96 (without C44), D45-D47) |
| Comorbidity 5: Moderate to severe renal disease | Denmark | *The National Patient Register.* Defined as primary diagnoses regardless of type of hospital contact registered before the start date for the country-specific rollout of the fourth vaccine dose (look-back 3 years). | Binary: yes/no (ICD-10 codes: I12, I13, N00–N05, N07, N11, N14, N17–N19, Q61) |
|  | Finland | *Care register for Health Care.* Defined as primary or secondary diagnoses prior to the start of the study period. | Binary: yes/no (ICD-10 codes: I12, I13, N00–N05, N07, N08, N11, N14, N18, N19, E102, E112, E142) |
|  | Sweden | *National Patient Register.* Defined as any recorded ICD-10 diagnosis during inpatient or outpatient contact and before first Covid-19 vaccination (look-back 3 years). | Binary: yes/no (ICD-10 codes: I12, I13, N00-N05, N07, N11, N14, N17-N19, Q61) |
| Covid-19 vaccine priority groups^b^ | Denmark | *The Danish Vaccination Register.* Defined as governmentally assigned Covid-19 vaccine priority groups, prioritised according to the risk of severe Covid-19 as well as whether being health and social care workers (last update 24 May 2021). | Categorical (3 levels): Severe Covid-19 risk group, healthcare personnel, others |
|  | Finland | *Register of Social Assistance.* Severe Covid-19 risk group was defined as vulnerable individuals in 24-hours care (binary status per 27 December 2021).  *Care register for Health Care (data since 1.1.2015), Special Reimbursement Register (data from 1.1.2018 to 27.12.2020) Prescription Centre database (data from 1.1.2018 to 27.12.2020.* Covid-19 risk group was defined on the basis of national vaccination recommendation (4). |  |
|  | Sweden | *Register on persons in nursing homes.* Severe Covid-19 risk group was defined as vulnerable individuals being residents at nursing homes (binary status as of 31 December 2020)  *The Longitudinal integrated database for health insurance and labour market studies.* Healthcare personnel defined as healthcare worker occupation status as of 31 October 2018 (binary). |  |
| Time since SARS-CoV-2 infection | Denmark | *The Danish Microbiology Database.* Defined as the date of any (last) registered positive PCR test for SARS-CoV-2 prior to the start date for the country-specific rollout of the fourth vaccine dose. | Categorical (for both adjustment and stratification; 4 levels): no previous infection, <6months, 6-12 months, >12 months |
|  | Finland | *National Infectious Diseases Register.* Defined as the date of any (last) registered positive PCR test for SARS-CoV-2 prior to the start date for the country-specific rollout of the fourth vaccine dose. |  |
|  | Sweden | *Register on surveillance of notifiable communicable diseases (SmiNet).* Defined as the date of any (last) registered positive PCR test for SARS-CoV-2 prior the start date for the country-specific rollout of the fourth vaccine dose. |  |

^a^Autoimmunity-related conditions include a range disorders such as inflammatory bowel diseases, diseases involving the blood, immune mechanism or endocrine systems, inflammatory rheumatic diseases, psoriasis, lupus erythematosus, multiple sclerosis; subject to country-specific definitions. The selected diagnosis codes to define comorbidities were country-specific, based on inputs from national experts and country specific registration practices as part of the general national surveillance purposes. This was done as we anticipated that country-specific definitions were likely better at identifying comorbidity-related risk groups within each country than a common set of code definitions. ^b^To account for the risk of severe Covid-19, we adjusted for targeted Covid-19 high-risk groups of severe Covid-19, specifically established for each country. In Denmark, the Covid-19 vaccine priority groups were governmentally assigned and individuals were prioritized according to the risk of severe infection (identified by the treating physicians) as well as whether being health or social care workers. In the remaining countries, the variable was constructed based on the identification of vulnerable individuals (as defined by those receiving nursing care or living in nursing homes) and whether being health or social care workers.

### Supplementary Figure S1. Density plots of the distribution of birth year and index date for the matched bivalent BA.4-5 or BA.1 boosted as ≥fourth vaccine dose vs unboosted comparisons in the immunocompromised population in the three Nordic countries.

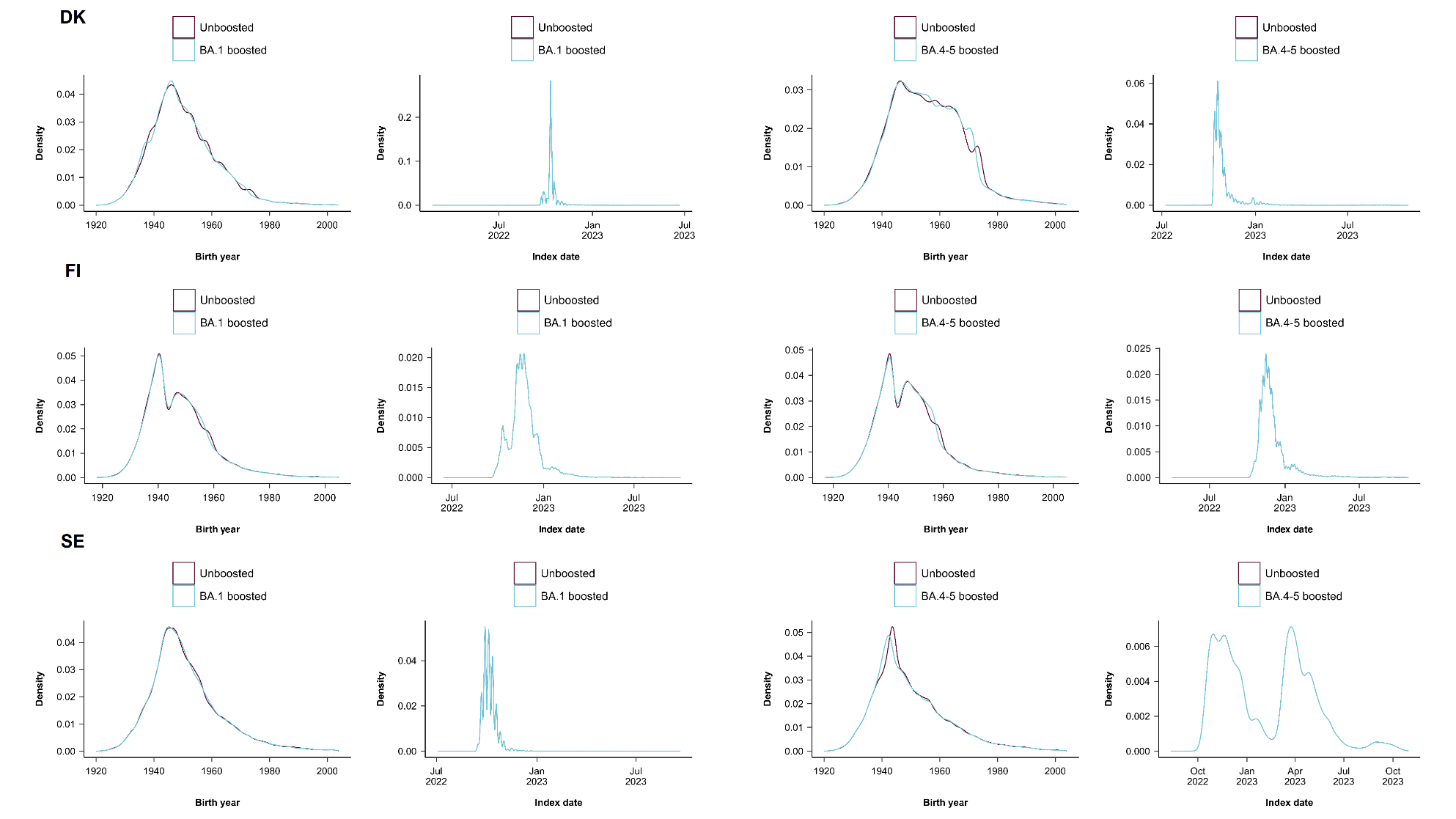

DK denotes Denmark, FI Finland, SE Sweden

### Supplementary Figure S2. Differences in proportions before and after matching of bivalent BA.4-5 or BA.1 boosted as ≥fourth vaccine dose vs unboosted in the immunocompromised population in the three Nordic countries.

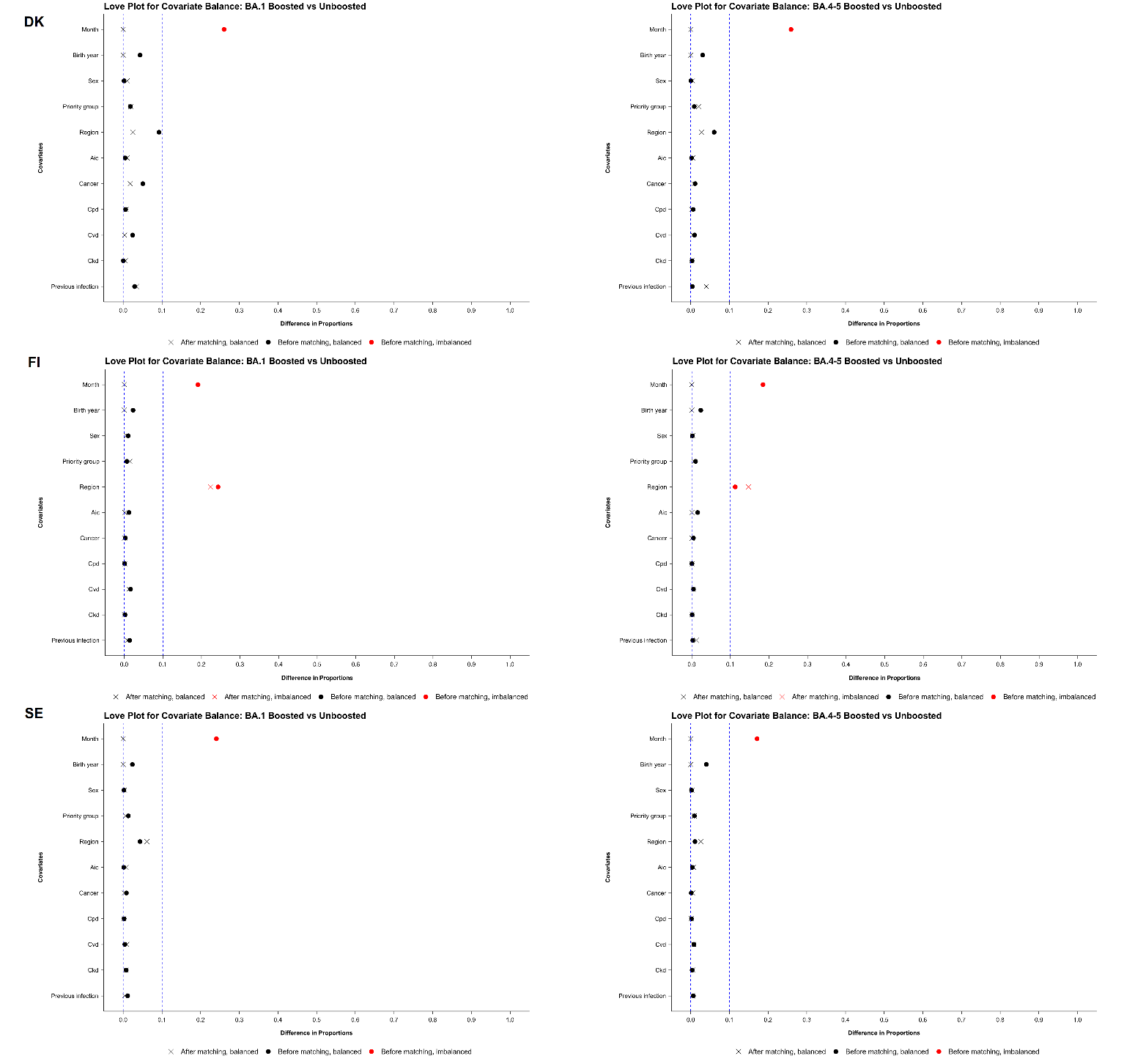

DK denotes Denmark, FI Finland, SE Sweden

### Supplementary References

1.	Embi PJ. Effectiveness of 2-dose vaccination with mRNA COVID-19 vaccines against COVID-19–associated hospitalizations among immunocompromised adults—nine states, January–September 2021. MMWR Morbidity and Mortality Weekly Report. 2021;70.

2.	Hughes K, Middleton DB, Nowalk MP, Balasubramani GK, Martin ET, Gaglani M, et al. Effectiveness of Influenza Vaccine for Preventing Laboratory-Confirmed Influenza Hospitalizations in Immunocompromised Adults. Clin Infect Dis. 2021;73(11):e4353-e60.

3.	Salo H, Lehtonen T, Auranen K, Baum U, Leino T. Predictors of hospitalisation and death due to SARS-CoV-2 infection in Finland: A population-based register study with implications to vaccinations. Vaccine. 2022;40(24):3345-55.

4.	Baum U, Poukka E, Leino T, Kilpi T, Nohynek H, Palmu AA. High vaccine effectiveness against severe COVID-19 in the elderly in Finland before and after the emergence of Omicron. BMC Infect Dis. 2022;22(1):816.
